# Temporal Trends in Utilization of Transvenous Lead Extractions among Patients with Cardiac Implantable Electronic Device Infections: Analyses of the Nationwide Japanese Registry of All Cardiac and Vascular Diseases-Diagnostic Procedure Combination data

**DOI:** 10.1101/2025.03.31.25324994

**Authors:** Toshihiro Nakamura, Yoko Sumita, Koshiro Kanaoka, Kimitake Imamura, Mitsuru Takami, Koji Fukuzawa

**Affiliations:** Department of Cardiovascular Medicine, National Cerebral and Cardiovascular Center, Suita, Japan; Department of Medical and Health Information Management, National Cerebral and Cardiovascular Center, Suita, Japan; Department of Cardiovascular Medicine, Nara Medical University, Kashihara, Japan; Section of Arrhythmia, Division of Cardiovascular Medicine, Department of Internal Medicine, Kobe University Graduate School of Medicine, Kobe, Japan; Division of Cardiovascular Medicine, Department of Internal Medicine, Kobe University Graduate School of Medicine, Kobe, Japan

**Keywords:** cardiac implantable electronic device, infection, lead extraction, antibiotics, complication

## Abstract

**Background:** Cardiac implantable electronic device (CIED) infections are a growing concern, leading to significant morbidity and mortality. International guidelines recommend a complete device and lead extraction for infected cases, yet real-world adherence to these recommendations remains suboptimal. The utilization of transvenous lead extractions (TLEs) in Japan has not been well characterized.

**Methods:** We conducted a retrospective observational study using the Japanese Registry of All Cardiac and Vascular Diseases diagnosis procedure combination/per diem payment system (JROAD-DPC) from 2015 to 2021. Patients diagnosed with CIED infections were identified based on the International Classification of Diseases, 10th revision codes. This study aimed to investigate the real-world utilization of TLEs in CIED infections, assess the trends over time, and identify the factors associated with the likelihood of undergoing a TLE and the procedural complications.

**Results:** Among 7,434 hospitalization records, only 38% underwent TLEs for CIED infections in the overall hospitals. However, the utilization of TLEs in certified TLE hospitals increased significantly from 54.0% in 2015 to 70.8% in 2021 (p-trend < 0.001). Independent predictors of reduced TLEs included a lower BMI, female sex, cerebrovascular disease, and dementia. Notably, 65.5% of patients in non-certified hospitals were discharged home, suggesting potential undertreatment.

**Conclusions:** While the TLE rate is increasing, a considerable proportion of patients with CIED infections do not undergo TLEs. Addressing barriers through physician education, improved referral systems, and adherence to guideline-directed management, especially in non-certified hospitals, is essential for optimizing the TLE rate.

**Clinical Perspectives:** *What Is New?:*

- This study was the first nationwide, population-based analysis to evaluate the actual rate and trends of transvenous lead extractions (TLEs) for cardiac implantable electronic device (CIED) infections in Japan.
- By rigorously identifying clinically indicated cases based on the current guidelines, it revealed a marked increase in TLE implementation, particularly in certified hospitals, and also highlighted a substantial undertreatment in non-certified centers.

*What Are the Clinical Implications?:*

- Despite increasing TLE rates, especially in certified hospitals, a significant proportion of patients with clinically indicated CIED infections do not undergo lead extractions.
- Identifying the predictors of undertreatment and reinforcing education and referral pathways in non-certified hospitals are essential steps to ensure a timely and appropriate guideline-directed management.

## 1. Introduction

Cardiac implantable electronic devices (CIEDs), including pacemakers and implantable cardioverter-defibrillators (ICDs), play a crucial role in managing various cardiac conditions, such as bradycardia, tachyarrhythmia, and heart failure. Despite their significant therapeutic benefits, CIED infections remain a serious and potentially fatal complication, with rising incidence over recent decades.^1, 2^ The infection risk increases with device longevity, with studies indicating that up to 11.7% of patients experience infections within 25 years of the implantation. Given the increasing prevalence of CIED implantations worldwide, effective infection management strategies, including lead extractions, are imperative.^3^

Transvenous lead extractions (TLEs) are the recommended standard of care for CIED-related infections, as endorsed by international guidelines.^3, 4^ Complete device and lead extractions are associated with a significantly improved survival compared to antibiotic therapy alone.^5^ Nevertheless, given the results of previous reports, the rate of lead extractions in cases of CIED infections was suboptimal.^5^ The underutilization of TLEs has been attributed to concerns over procedural risks, lack of expertise, and healthcare system inefficiencies.^5^ Previous reports have highlighted the notably low utilization of TLEs for CIED infections.^6, 7^ However, we believe that these findings differ substantially from the actual clinical practice.

The Japanese Lead Extraction Registry (J-LEX) provides essential data on TLE practice. According to the J-LEX report, 62.8% of lead extraction procedures were performed for infection-related indications, with a high clinical success rate of 98.1%.^8^ While J-LEX data provide insights into the number of lead extraction procedures performed, the actual incidence of CIED infections requiring lead extractions in Japan remains unknown, making it difficult to assess the true rate of appropriate TLE implementations.

This study aimed to elucidate the actual rate of TLEs for CIED infections, the current status of undertreatment, and the improvement trends using a national registry database.

## 2. Methods

### Data source

We conducted a multicenter retrospective observational study using the Japanese Registry of All Cardiac and Vascular Diseases diagnosis procedure combination/per diem payment system (JROAD-DPC). More than 60% of admissions in cardiovascular departments in the Japanese Circulation Society-certified cardiology training hospitals in Japan are included in the database. The JROAD-DPC is a nationwide claim-based database based on data from the Japanese DPC/Per Diem Payment System in hospitals participating in the JROAD survey.^9^ The JROAD database has been described in detail previously.^10, 11^ In brief, all teaching hospitals with cardiovascular beds participate in the JROAD. Hospital doctors must record all admission and discharge data for each patient and are obliged to guarantee their accuracy.

The JROAD-DPC database consists of the following information for each patient discharged: age; sex; height; weight; primary diagnoses/comorbidities/conditions arising after admission based on the International Classification of Diseases, 10th revision (ICD-10) codes; the Charlson comorbidity index; diagnostic and therapeutic procedures; and discharge status.

### Study population

From April 2015 to March 2022, patients diagnosed with CIED infections were identified in the JROAD database. Patients were included in this study if they had ICD-10 codes T82.7 or T82.8 with any of the following: “main diagnosis,” “admission-precipitating diagnosis,” and “most resource-consuming diagnosis” and “second-most resource-consuming diagnosis”. Additionally, patients were required to have an ICD-10 code corresponding to an infectious disease listed in the Supplemental materials (see ***Table S1***). The recorded diagnoses in the medical texts were meticulously reviewed to exclude ambiguous cases, such as suspected infections and hospitalizations for reasons unrelated to the treatment of CIED infections. Patients who received antibiotics for fewer than three days were excluded, as such cases were unlikely to represent CIED infections, given the latest guidelines recommending a minimum of 2 to 4 weeks of antibiotic therapy for CIED infections.^3^ We also considered antibiotics that are often administered within three days in clinical practice, even in cases of newly implanted devices and generator exchanges. Subsequently, records with readmissions were excluded. The baseline characteristics, clinical presentation, management, and in-hospital outcomes, including death, length of hospitalization, medical cost, and discharge locations, were analyzed. The procedure codes used to identify treatment methods and ICD-10 codes used to identify comorbidities are summarized in the Supplemental materials (see ***Table S2, S3***). We evaluated the annual TLE rate across the overall hospitals and specifically in hospitals certified by the Japanese Heart Rhythm Society (JHRS) to perform TLEs. The criteria for certified TLE hospitals require the presence of a full-time cardiologist with expertise in lead extraction procedures as well as a cardiovascular surgeon. In Japan, TLEs are exclusively performed at certified hospitals. If a hospitalization record had both procedure codes for a TLE and surgical lead extraction, it was classified under the TLE group. This classification was based on the assumption that the TLE would have been attempted first, with a transition to surgical lead extraction only if the TLE was unsuccessful or if a severe complication occurred. To assess the real-world treatment practices in non-certified hospitals, the discharge location was also examined. Patients requiring lead extractions at non-certified hospitals should have been transferred to specialized hospitals. Conversely, the patients discharged home may have included those who were classified as having received undertreatment, which meant a lead extraction was not performed despite being indicated.

Furthermore, the occurrence and clinical outcomes of TLE-associated complications (vascular complications, hematoma/hemorrhage, cardiac tamponade, and pneumothorax/hemothorax, [listed in Supplementary materials, ***Table S4***]) were reviewed.

### Statistical Analysis

Categorical data are given as the frequency (percentage). Continuous variables were expressed as medians and interquartile ranges (IQRs). The Mann–Whitney U test was performed to compare the two groups. The chi-square test was used to compare the categorical variables. At first, to assess the clinical predictors of the utilization of lead extractions, a univariate Cox proportional analysis was performed. Sequentially, variables with a P<0.10 in the univariate analysis were included in the multivariate analysis, and the hazard ratios (HR) and 95% confidence interval (CI) were calculated. All statistical comparisons were two-sided, and a p < .05 was considered significant. All statistical analyses were performed with STATA 17.0 software (StataCorp LP, College Station, TX, USA).

## 3. Results

### Baseline Characteristics

A total of 7,434 hospitalization records from 752 hospitals with a primary diagnosis of a CIED infection from April 2015 to March 2022, were extracted from the database. The patient selection flowchart is shown in **Figure 1**. After excluding non-eligible patients, 37.9% (n = 2,180) and 3.3% (n = 191) were managed with TLEs and surgical extractions, respectively.

**Figure 1.**
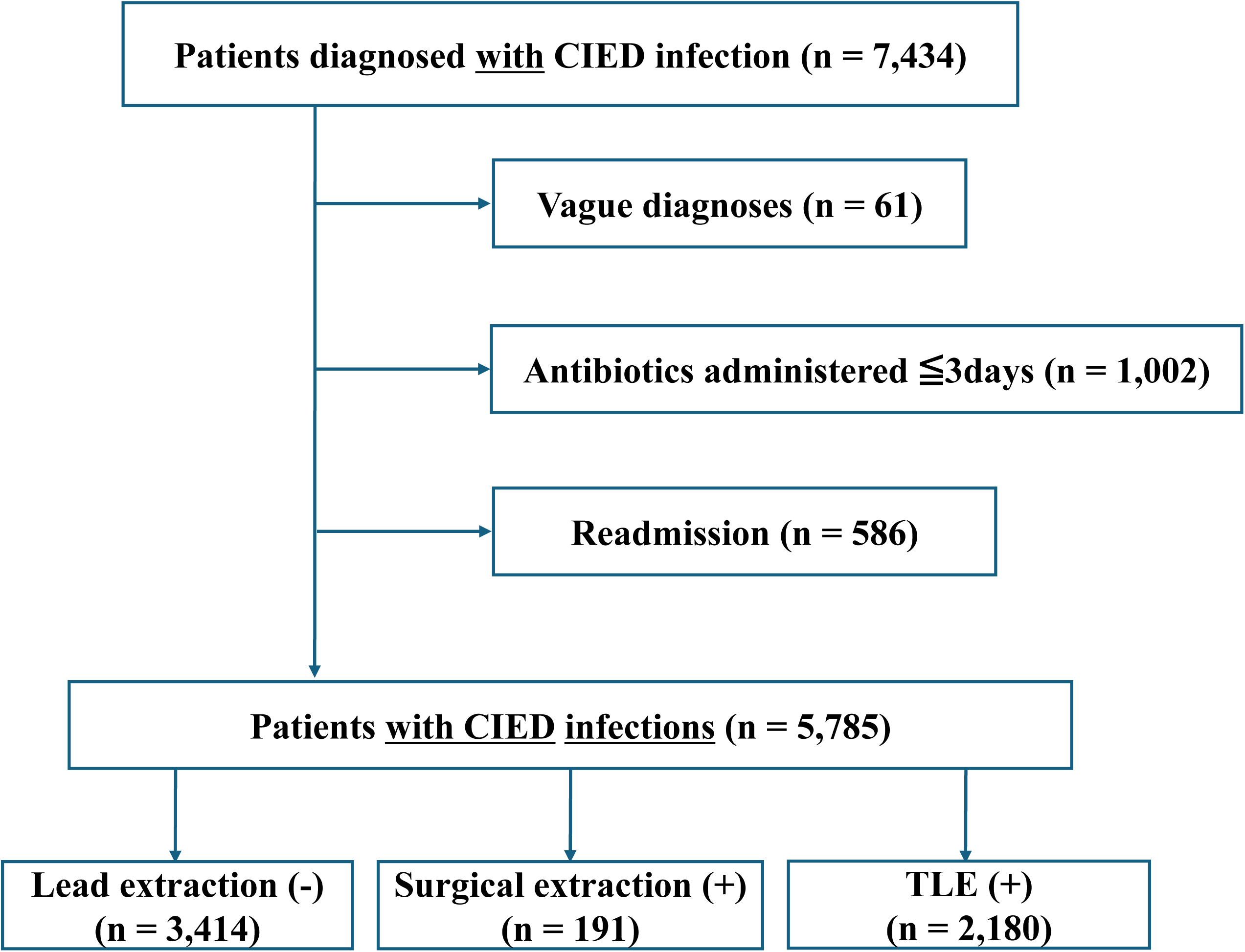
Study flowchart. CIED, cardiac implantable electronic devices; TLE, transvenous lead extraction

Baseline characteristics for the overall population and a comparison of the patients who underwent TLEs and those who did not are shown in **Table 1**. The overall population had a median age of 79 (71-85) years and consisted of 31.1% female patients.

**Table 1.** Patient characteristics in the overall population with or without TLEs.

|  | Overall<br>n = 5,785 | TLE (-)<br>n = 3,605 | TLE (+)<br>n = 2,180 | P value |
| --- | --- | --- | --- | --- |
| Age, years | 79.0 (71.0 - 85.0) | 79.0 (71.5-86.0) | 78.0 (70.0-84.0) | <0.001 |
| Female | 1,799 (31.1) | 1,231 (34.1) | 568 (26.1) | <0.001 |
| Body mass index, kg/m<sup>2</sup> | 22.1 (19.7, 24.8) | 22.0 (19.5-24.7) | 22.4 (20.1-24.9) | <0.001 |
| Congestive heart failure | 2,361 (40.8) | 1,513 (42.0) | 848 (38.9) | 0.021 |
| Myocardial infarction | 255 (4.4) | 157 (4.4) | 98 (4.5) | 0.80 |
| Renal disease | 569 (9.8) | 378 (10.5) | 191 (8.8) | 0.033 |
| Diabetes mellitus | 1,326 (22.9) | 875 (24.3) | 451 (20.7) | 0.002 |
| Cerebral vascular disease | 279 (4.8) | 218 (5.8) | 61 (2.9) | <0.001 |
| Cancer | 203 (3.5) | 154 (4.3) | 49 (2.2) | <0.001 |
| Dementia | 1,340 (23.2) | 931 (25.8) | 409 (18.8) | <0.001 |
| Hospitalization days | 23.0 (14.0-36.0) | 20.0 (12.0-34.0) | 28.0 (19.0-39.0) | <0.001 |
| Charlson comorbidity index score |  |  |  | <0.001 |
| 0 | 1,183 (31) | 974 (27) | 807 (37) |  |
| 1 | 1,197 (35) | 1,261 (35) | 741 (34) |  |
| ≥2 | 1,087 (34) | 1,370 (38) | 632 (29) |  |
| Direct cost, US dollar | 19190.8 (7717.2-33320.0) | 11526.6 (4998.8-21953.9) | 31645.7 (22026.8-41760.8) | <0.001 |
| Died | 167 (2.9) | 119 (3.3) | 48 (2.2) | 0.016 |
| Discharge location |  |  |  | <0.001 |
| Home | 3,869 (66.9) | 2,390 (66.3) | 1,479 (67.8) |  |
| Transfer | 1,507 (26.1) | 892 (24.7) | 615 (28.2) |  |
| Nursing home | 220 (3.8) | 187 (5.2) | 33 (1.5) |  |
| Other | 189 (3.3) | 136 (3.8) | 53 (2.4) |  |

TLE, transvenous lead extraction
Values are the n (%) or median and interquartile ranges

Compared with the patients who did not receive TLEs, the patients who received a TLE were younger (median age 78 [70-84] years vs. 79 [71.5-86] years; P < 0.001), less likely to be female (568 [26.1%] vs. 1,1231 [34.1%]; P < 0.001) and had a higher body mass index (BMI) (22.4 [20.1-24.9] vs. 22.1 [19.7-31.1], P < 0.001). Patients who did not undergo a TLE had a higher prevalence of comorbidities, including congestive heart failure, renal disease, diabetes mellitus, cerebrovascular disease, cancer, and dementia. A similar trend was observed in certified TLE hospitals, where TLEs were often not performed in patients with substantial comorbidities or dementia (**Table 2**). The baseline characteristics of the patients with or without lead extractions (TLEs and surgical extractions) in the overall and certified TLE hospitals are shown in ***Table S5, S6***.

**Table 2.** Patient characteristics in certified TLE hospitals with or without TLEs.

|  | Overall<br>n = 3,467 | TLE (-)<br>n = 1,287 | TLE (+)<br>n = 2,180 | P value |
| --- | --- | --- | --- | --- |
| Age, years | 78.0 (69.0-84.0) | 77.0 (68.0-85.0) | 78.0 (70.0-84.0) | 0.63 |
| Female | 986 (28.4) | 418 (32.5) | 568 (26.1) | <0.001 |
| Body mass index, kg/m<sup>2</sup> | 22.3 (19.8-24.8) | 22.0 (19.5-24.6) | 22.4 (20.1-24.9) | 0.008 |
| Congestive heart failure | 1,406 (40.6) | 508 (43.8) | 898 (38.9) | 0.006 |
| Myocardial infarction | 160 (4.6) | 62 (4.8) | 98 (4.5) | 0.66 |
| Renal disease | 334 (9.6) | 143 (11.1) | 191 (8.8) | 0.023 |
| Diabetes mellitus | 737 (21.3) | 286 (22.2) | 451 (20.7) | 0.47 |
| Cerebral vascular disease | 111 (3.2) | 50 (3.9) | 61 (2.8) | 0.079 |
| Cancer | 96 (2.8) | 47 (3.7) | 49 (2.2) | 0.005 |
| Dementia | 732 (21.1) | 323 (25.1) | 409 (18.8) | <0.001 |
| Hospitalization days | 26.0 (16.0-38.0) | 19.0 (11.0-33.0) | 29.0 (19.0-41.0) | <0.001 |
| Charlson comorbidity index score |  |  |  | <0.001 |
| 0 | 1,183 (34) | 360 (28) | 806 (37) |  |
| 1 | 1,197 (35) | 450 (35) | 741 (34) |  |
| ≥2 | 1,087 (31) | 476 (37) | 632 (29) |  |
| Direct cost, US dollar | 26309.6 (15039.5-38764.0) | 12906.4 (5233.6-27536.9) | 32165.7 (22026.8-41760.8) | <0.001 |
| Died | 96 (2.8) | 111 (3.3) | 56 (2.4) | 0.047 |
| Discharge location |  |  |  | <0.001 |
| Home | 2,350 (67.8) | 871 (67.7) | 1,479 (67.8) |  |
| Transfer | 925 (26.7) | 310 (24.1) | 615 (28.2) |  |
| Nursing home | 85 (2.5) | 52 (4.0) | 33 (1.5) |  |
| Other | 107 (3.1) | 54 (4.2) | 53 (2.4) |  |

TLE, transvenous lead extraction
Values are the n (%) or median and interquartile ranges

The trends in the proportion of patients with CIED infections undergoing TLEs are shown in **Figures 2A and 2B**. Between 2015 and 2021, there was a significant increase in the proportion of patients undergoing TLEs in both the overall population (increasing from 30.6% in 2015 to 44.0% in 2021, P trend < 0.001) and certified TLE hospitals (increasing from 54.0% in 2015 to 70.8% in 2021, P trend < 0.001). Moreover, the trends in the proportion of patients with CIED infections undergoing lead extractions (TLEs and surgical extractions) are shown in **Figures S1A and S1B**. As with TLEs, there was a significant increase in the proportion of patients undergoing lead extractions in both the overall population (increasing from 35.3% in 2015 to 46.4% in 2021, P trend < 0.001) and certified TLE hospitals (increasing from 59.6% in 2015 to 73.1% in 2021, P trend < 0.001).

**Figure 2.**
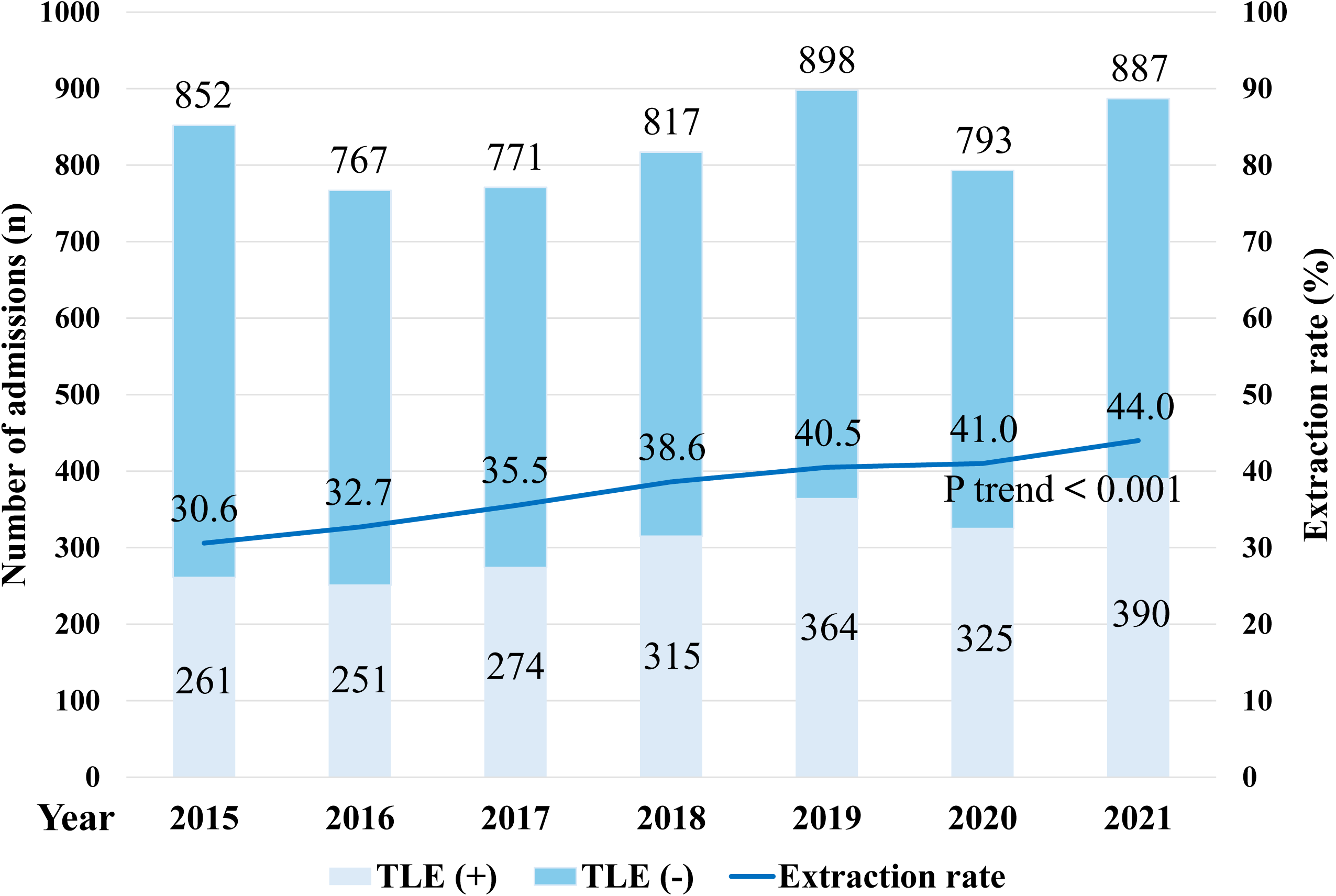
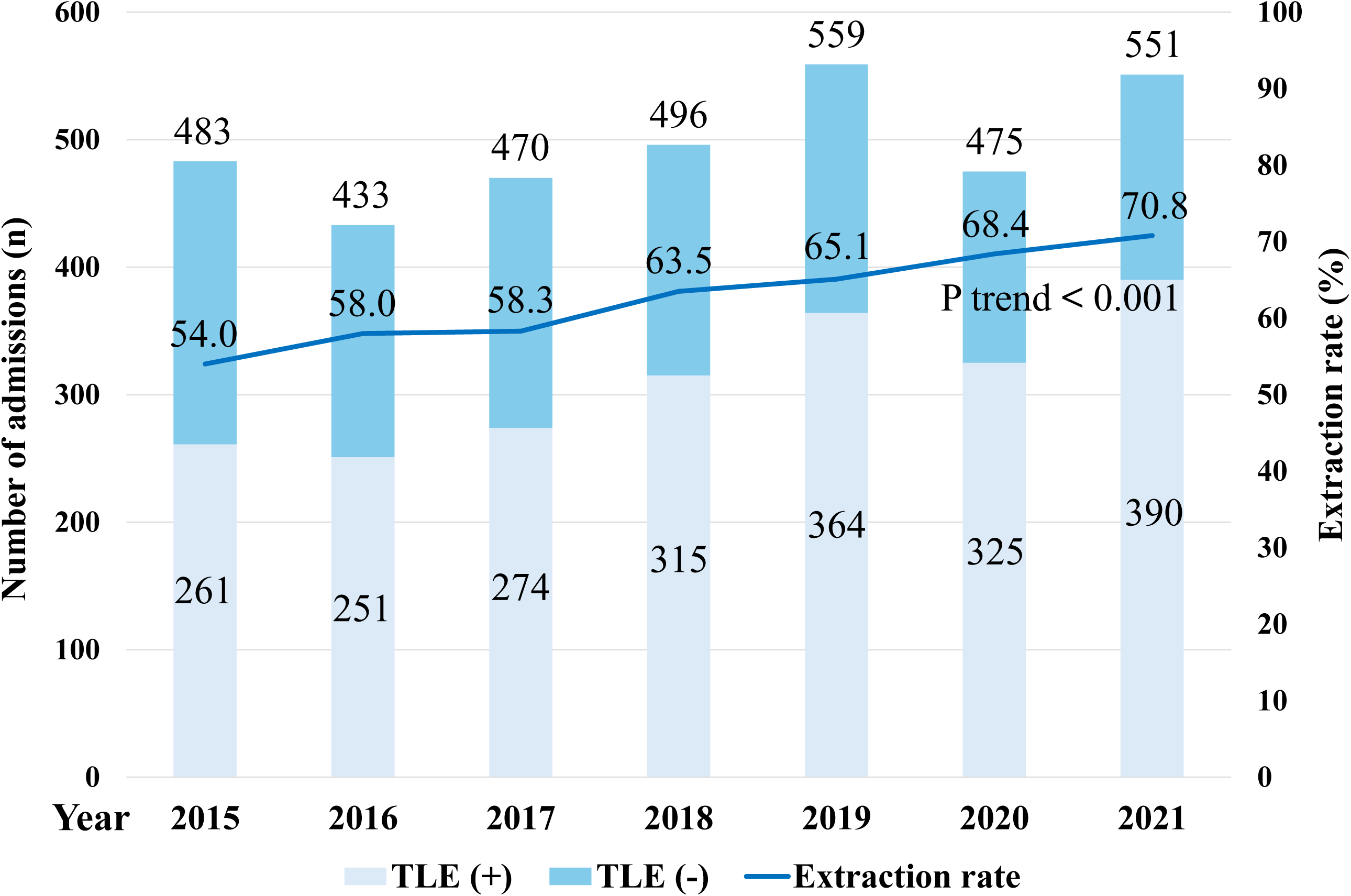
The proportion of admissions with cardiac implantable electronic device infections undergoing TLEs in the overall (**A**) and certified TLE hospitals (**B**), as recorded in the Japanese Registry of All Cardiac and Vascular Diseases database from 2015 to 2021. TLE, transvenous lead extraction

The proportion of the discharge locations in the non-certified TLE hospitals is shown in **Figure 3**. Of note, 65.5% of the CIED infection patients were discharged to their homes, and only 31.5% of the patients were transferred even in the most recent analysis year of 2021.

**Figure 3.**
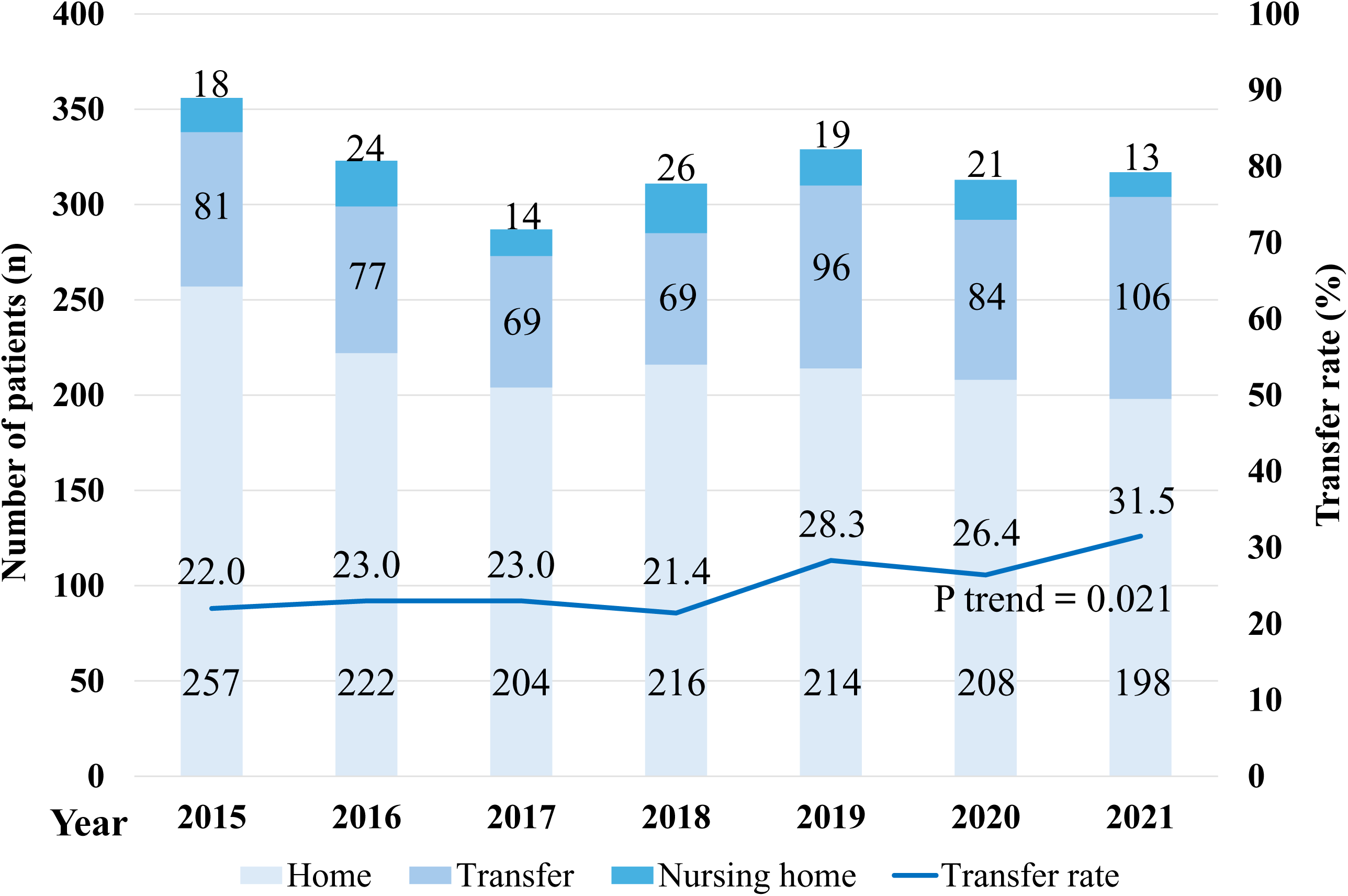
Discharge locations in the non-certified TLE hospitals. TLE, transvenous lead extraction

#### Predictors of Lead Extractions

Multivariable analysis examining the factors independently associated with the TLE management are presented in **Table 3**. The BMI (for each 1 increase; OR: 1.01; 95% CI: 1.01-1.03) was independently associated with the TLE management. In contrast, factors associated with a lower likelihood of a lead extraction included the presence of a female sex (OR: 0.74; 95% CI: 0.67-0.82), cerebral vascular disease (OR: 0.65; 95% CI: 0.49-0.85), and dementia (OR: 0.73; 95% CI: 0.58-0.98). An additional multivariable analysis examining the predictors associated with any form of lead extraction (TLEs or surgical extractions) is provided in ***Table S7***. The findings were consistent with those obtained for a TLE alone.

**Table 3.** Predictors of TLEs among the patients with CIED infections.

|  | Univariable |  | Multivariable |  |
| --- | --- | --- | --- | --- |
|  | OR (95% CI) | P value | OR (95% CI) | P value |
| Age | 0.99 (0.99-0.99) | <0.001 | 0.99 (0.95-1.02) | 0.369 |
| Female | 0.72 (0.66-0.80) | <0.001 | 0.74 (0.67-0.82) | <0.001 |
| Body mass index (1 decrease) | 0.98 (0.97-0.99) | <0.001 | 0.98 (0.97-0.99) | 0.005 |
| Congestive heart failure | 0.75 (0.69-0.82) | <0.001 | 0.92 (0.82-1.03) | 0.143 |
| Myocardial infarction | 0.94 (0.77-1.16) | 0.580 |  |  |
| Renal disease | 0.67 (0.58-0.78) | <0.001 | 1.07 (0.85-1.34) | 0.561 |
| Diabetes mellitus | 0.82 (0.74-0.91) | <0.001 | 1.08 (0.95-1.23) | 0.248 |
| Cerebral vascular disease | 0.50 (0.38-0.64) | <0.001 | 0.65 (0.50-0.85) | 0.002 |
| Cancer | 0.47 (0.35-0.62) | <0.001 | 0.70 (0.50-0.97) | 0.031 |
| Dementia | 0.55 (0.43-0.70) | <0.001 | 0.76 (0.59-0.98) | 0.035 |
| Charlson comorbidity index score (1 increase) | 0.78 (0.75-0.81) | <0.001 | 0.80 (0.74-0.86) | <0.001 |

CIED, cardiac implantable electronic devices; TLE, transvenous lead extraction

#### Complications in TLE Procedures

Among the patients managed with a TLE, procedural complications were identified in 2.8%, which included superior vena cava injury/innominate vein injury/other vascular complications in 0.9%, cardiac perforation/tamponade in 1.5%, pulmonary embolism in 0.4%, and pneumothorax/hemothorax in 1.8%. The percentage of all patients managed with TLEs who died and experienced a TLE-associated procedural complication was 0.2%. The patient characteristics with and without complications in the TLE procedures are summarized in **Table 4**. Patients with complications during TLE procedures had a higher incidence of cancer (6 [10%] vs. 43 [2.0%]; P < 0.001), longer hospitalization days (median 41.0 [26.5-58.0] days vs. 28.0 [19.0-39.0] days; P < 0.001), higher direct cost (median 32195.2 [22408.8-44023.1] US dollars vs. 10495.2 [4924.7-22453.4] US dollars; P < 0.001), and higher mortality rate (4 [6.7%] vs. 44 [2.1%]; P < 0.001).

**Table 4.** Patient characteristics with and without complications during the TLE procedures.

|  | Complication (-)<br>n = 2,120 | Complication (+)<br>n = 60 | P value |
| --- | --- | --- | --- |
| Age, years | 78.0 (70.0-84.0) | 76.0 (68.0-84.5) | 0.56 |
| Female | 552 (26.0) | 16 (26.7) | 0.91 |
| Body mass index, kg/m<sup>2</sup> | 22.4 (20.1-24.9) | 21.8 (18.7-25.5) | 0.46 |
| Congestive heart failure | 825 (38.9) | 23 (38.3) | 0.93 |
| Myocardial infarction | 96 (4.5) | 2 (3.3) | 0.66 |
| Renal disease | 184 (8.7) | 7 (11.7) | 0.42 |
| Diabetes mellitus | 437 (20.6) | 14 (23.3) | 0.61 |
| Cerebral vascular disease | 60 (2.8) | 1 (1.7) | 0.59 |
| Cancer | 43 (2.0) | 6 (10.0) | <0.001 |
| Dementia | 390 (18.4) | 19 (31.7) | 0.009 |
| Hospitalization days | 28.0 (19.0-39.0) | 41.0 (26.5-58.0) | <0.001 |
| Charlson comorbidity index score |  |  | 0.26 |
| 0 | 721 (34) | 17 (28) |  |
| 1 | 742 (35) | 21 (35) |  |
| ≥2 | 657 (31) | 22 (37) |  |
| Direct cost, US dollar | 10495.2 (4924.7-22453.4) | 32195.2 (22408.8-44023.1) | <0.001 |
| Died | 44 (2.1) | 4 (6.7) | 0.017 |
| Discharge location |  |  | 0.11 |
| Home | 1444 (67.3) | 35 (58.3) |  |
| Transfer | 595 (24.2) | 20 (33.3) |  |
| Nursing home | 32 (4.3) | 1 (1.7) |  |
| Other | 4 (0.2) | 0 (0) |  |

TLE, transvenous lead extraction
Values are the n (%) or median and interquartile ranges

## 4. Discussion

### Main findings

In this study of a contemporary largest population-based study comparing the characteristics, management, and outcomes of Japanese patients with CIED infections who underwent TLEs to those who did not, we identified several key findings (**Central Illustration**). First, only 39% of patients with CIEDs underwent TLEs despite the diagnosis of CIED infections, even though there were low overall rates of TLE-related complications at 2.8%. The proportion of patients undergoing TLE management in certified TLE hospitals increased significantly from 54.0% in 2015 to 70.8% in 2021. Second, a lower BMI, female sex, cerebral vascular disease, and dementia were independently associated with a decreased likelihood of a lead extraction as part of the CIED infection management.

#### Identification of Definitive CIED infections requiring TLEs

Previous reports have defined CIED infections solely based on the presence of an infectious disease diagnosis or a history of antibiotic therapy.^6, 7, 12^ However, such criteria may include superficial infections that do not warrant a lead extraction according to the latest guidelines, as well as cases of new implantations or generator exchanges in which antibiotics were administered for infection prophylaxis. In this study, we aimed to identify true CIED infections that necessitated TLEs as accurately as possible and calculated the actual lead extraction rate. Moreover, TLEs are only performed at hospitals certified by the JHRS. Consequently, the lead extraction rate for CIED infections at certified TLE hospitals has increased up to 70% in the most recent analysis year of 2021. It is unlikely that our method of identification underestimatd the incidence of CIED infections; however, it remains possible that the TLE rate was insufficient. In clinical practice, TLEs may not be performed in patients with dementia and multiple comorbidities. Some patients diagnosed with CIED infections may not require TLEs.

In this study, we excluded patients who underwent TLEs during a readmission. as a delayed lead extraction is associated with increased complications and mortality. A study by Lin et al. found that patients who experienced delays in lead extractions had a two-fold increase in mortality compared to those who underwent an early extraction.^13^

The risk of systemic dissemination of an infection, particularly lead-associated endocarditis, further underscores the necessity of a timely lead extraction.^14^ These findings emphasized the importance of a prompt diagnosis and guideline-directed management. Although including these patients would have increased the overall TLE rate, we believe that an early diagnosis and intervention are essential; therefore, the TLE rate was calculated based solely on the initial admission data.

#### Underutilization of TLEs in CIED Infections

Despite the overwhelming evidence supporting lead extractions in CIED infections,^15, 16^ the number of patients who have received this life-saving intervention is not sufficient. Greenspon et al. reported that as recently as 2018, 36% of patients with CIED infections did not undergo a complete system extraction.^5^ The consequences of non-adherence to the guidelines are profound, with increased mortality, recurrent infections, and prolonged hospitalizations.^17^

There have been several barriers contributing to this underutilization. First, many physicians perceive TLEs as a high-risk procedure, despite data showing a high success rate (>95%) in experienced centers.^18^ It should be noted that the overall procedural complication rates related to TLEs in the present cohort was low at 2.8%, which is comparable to the registry data from experienced centers showing TLE-related complication rates of 1.4% to 1.7%.^19, 20^ Second, non-electrophysiology specialists may lack familiarity with the latest guideline recommendations.^21^ Consequently, the initial treating physician may not recognize the need for an extraction, which leads to delaying a specialist referral.^18^ In addition, limited access to experienced operators in some institutions further delays an appropriate intervention.^22^

Misclassification of superficial infections may also lead to undertreatment, as complete extraction is not required in true superficial or incisional infections that do not involve the device. However, failure to accurately diagnose deeper infections risks missing clinically indicated extractions. A recent survey indicated that a knowledge gap persists between physicians who perform TLE and those who do not in the management of CIED infections.^23^ During the survey, approximately 10% of cardiologists and 40% of non-cardiologist physicians opted for a partial device removal in cases of CIED pocket infections.^23^

To address these issues, regional referral networks and multidisciplinary teams involving electrophysiologists, infectious disease specialists, and cardiovascular surgeons should be established to guide timely and evidence-based decision-making.

In this study, no increase was observed in the number of patient transfers from non-certified hospitals. Although the JROAD data do not include information on transfer destinations, it can be assumed that there has been no apparent rise in transfers specifically for TLEs from non-certified institutions. In fact, 65.5% of the patients with CIED patients admitted to non-certified hospitals were discharged to home. It is true that a complete extraction is not required for superficial or incisional infections at the device pocket if an infection does not involve the device.^24^ However, a misdiagnosis of a superficial infection means the potential risk of undertreatment, meaning that a lead extraction was not performed despite being clinically indicated. To bridge the gap between the guideline recommendations and real-world practice, a multifaceted approach is needed, focusing on physician education, especially in non-certified TLE hospitals, and could lead to higher TLE rates.

#### Study Limitations

There were several limitations to the present study. First, because this study was a retrospective analysis of claim-based data, there may have been risks of systemic or non-systemic coding errors or insufficient data collection. However, the validity of the DPC database was generally high, especially for primary diagnoses and procedure records.^25^

Second, since individuals could not be identified across the different hospitals in the J-ROAD study, there was a possibility that some cases were transferred to another hospital for a lead extraction or subsequently transferred for ongoing medical treatment following a lead extraction. Third, a possible overestimation of the actual infected population may have contributed to the observed low rates of lead extractions. Our focus was on avoiding an overestimation of the TLE rate that could result from an underestimation of true CIED infections. At the very least, we are confident that the actual TLE rate is not lower than this. Fourth, the actual lead removal rate may be higher than reported, as it was challenging to exclude patients with superficial infections that did not require a lead removal solely based on the disease classification.

## 5. Conclusion

In contemporary clinical practice in Japan, the TLE rate has increased from 38% in the overall hospitals to 70% in the certified TLE hospitals. However, 30% of patients with CIED infections still did not undergo a TLE, despite the low rate of procedural complications. Patients with a lower BMI, female sex, cerebrovascular disease, and dementia were less likely to receive a TLE. Enhancing education on the management of CIED infections in non-certified hospitals may contribute to an increased implementation of TLEs.

## Data Availability

Not applicable.

## Acknowledgments

The authors thank Mr. John Martin for his linguistic assistance.

## Data Availability Statement

Not applicable.

## List of Abbreviations

| **List of Abbreviations** |  |
| --- | --- |
| CIED | Cardiac implantable electronic device |
| ICD | Implantable cardioverter–defibrillator |
| ICD-10 | International Classification of Diseases, 10th revision |
| IQR | interquartile range |
| JHRS | Japanese Heart Rhythm Society |
| J-LEX | Japanese Lead Extraction Registry |
| JROAD-DPC | Japanese Registry of All Cardiac and Vascular Diseases diagnosis<br>procedure combination/per diem payment system |
| TLE | Transvenous lead extraction |

## Figure Legends

### Central Illustration

National Utilization of Transvenous Lead Extraction among Patients with Cardiac Implantable Electronic Device Infections

BMI, body mass index; CVD, cerebral vascular disease; TLE, transvenous lead extraction

